# Use of compressed sensing to expedite high-throughput diagnostic testing for COVID-19 and beyond

**DOI:** 10.1101/2021.08.09.21261669

**Authors:** Kody A. Waldstein, Jirong Yi, Michael Myung Cho, Raghu Mudumbai, Xiaodong Wu, Steven M. Varga, Weiyu Xu

## Abstract

The rapid spread of SARS-CoV-2 has placed a significant burden on public health systems to provide rapid and accurate diagnostic testing highlighting the critical need for innovative testing approaches for future pandemics. In this study, we present a novel sample pooling procedure based on compressed sensing theory to accurately identify virally infected patients at high prevalence rates utilizing an innovative viral RNA extraction process to minimize sample dilution. At prevalence rates ranging from 0-14.3%, the number of tests required to identify the infection status of all patients was reduced by 75.6% as compared to conventional testing in primary human SARS-CoV-2 nasopharyngeal swabs and a coronavirus model system. Additionally, our modified pooling and RNA extraction process minimized sample dilution which remained constant as pool sizes increased. Our use of compressed sensing can be adapted to a wide variety of diagnostic testing applications to increase throughput for routine laboratory testing as well as a means to increase testing throughput to combat future pandemics.

**Graphical Abstract:** 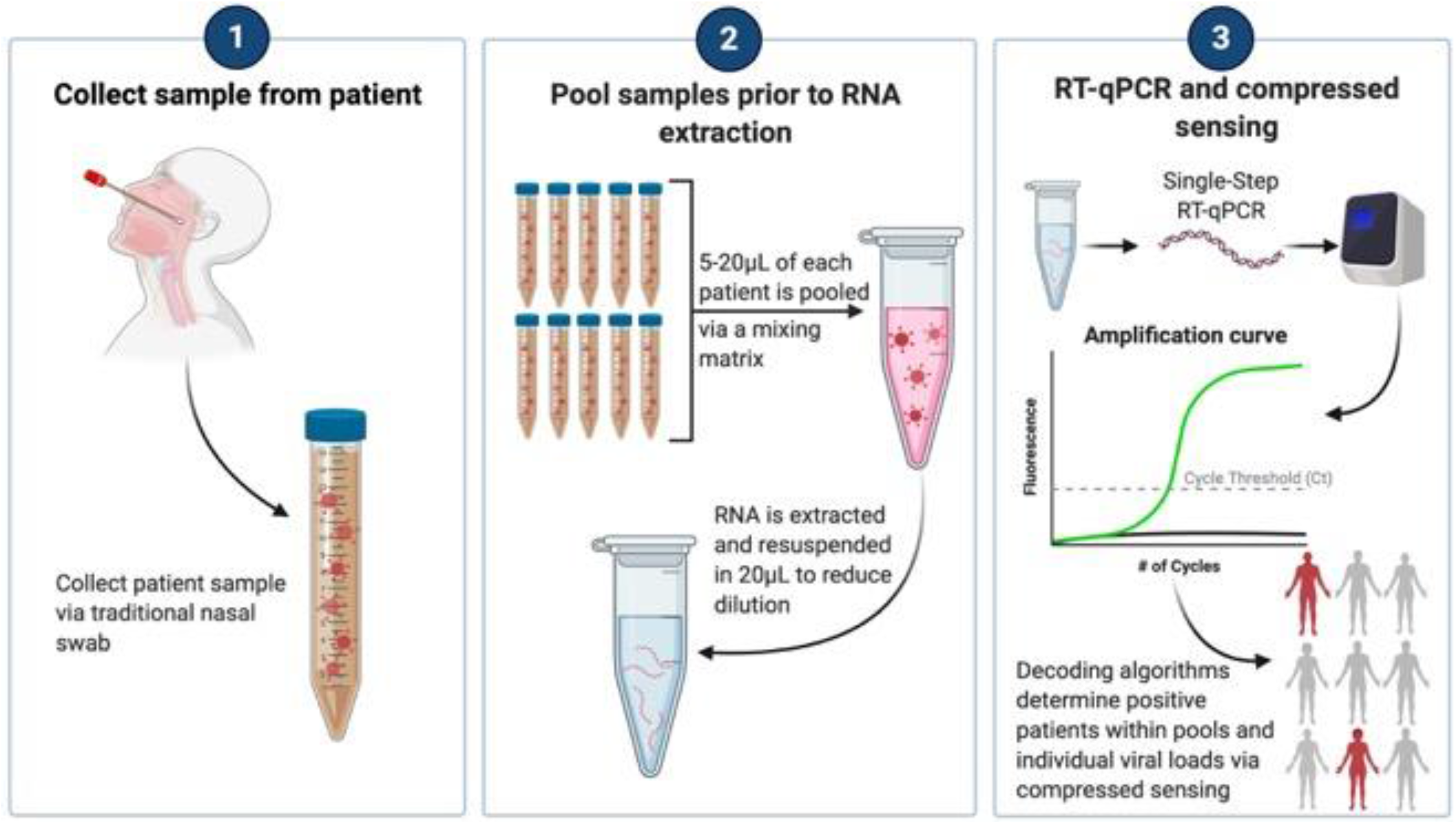

## Introduction

The rapid community spread of SARS-CoV-2 has placed a significant burden on diagnostic testing and public health to provide fast and accurate testing strategies. The number of COVID-19 tests being performed each day has increased 8-fold since testing reagents became widely available with an average of over 1.5-2 million COVID-19 quantitative reverse transcription polymerase chain reaction (qRT-PCR) tests performed by day in the United States alone (1-3). Additionally, multiple new and more infectious variants of COVID-19 have emerged worldwide harboring genetic mutations significant enough to evade recognition by host antibodies causing some concern for current vaccine formulations (4-8). Testing and screening remains an imperative safeguard to minimize spread, thus the development of innovative strategies and techniques to increase testing capacity without reducing the accuracy and efficacy of testing is crucial.

A traditional method to increase testing capacity is by pooling samples as opposed to conducting individualized testing, known as “group testing” (9-11). The principle is simple, if the prevalence rate is low within the population, the majority of samples will inevitably test negative. In this scenario, a single negative result indicates that all patients within that pool are also negative. However, the ability to accurately test using this method diminishes quickly as the prevalence rate increases (12-15). Current CDC guidelines require subsequent individual testing of all patients within a pool if the pool is positive (16). Worldwide SARS-CoV-2 prevalence rates continue to be >10% with a worldwide estimate of ∼30% (17). These rates are well beyond the capacity of traditional pooling methods as many pools will be positive requiring additional individual testing and inevitably increasing the number of tests required. More sophisticated pooling efforts have arisen during the pandemic though the testing models’ accuracy and effectiveness falls apart rapidly as the prevalence rate rises and are thus not viable options for the current and future pandemics (13, 16, 18, 19).

In this study, we present a novel and innovative pooling protocol which utilizes mathematically-derived mixing matrices and decoding algorithms to accurately identify positive patients within pools using the CDC-approved range of positive Ct values at high prevalence rates. Additionally, we propose a new approach based on compressed sensing theory for detection of viral load using pooled sample testing (20-22). We also employ a modified RNA extraction process in which the patient swab samples are pooled prior to RNA extraction allowing the sample to be concentrated thus minimizing sample dilution. This modified approach has shown high accuracy and reproducibility at prevalence rates over 10% with large sample sizes using an experimental mouse coronavirus, mouse hepatitis virus strain 1 (MHV-1) as well as human COVID-19 patient samples.

### Problem Formulation

Notations: We use [*N*] to denote the set {1, 2,…, *N*}, and 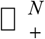 to denote the set [0, +∞)^*N*^. We denote by Pr(*E*) the probability of an event *E*, and use round(*x*) to round *x* to the closest integer. The *j* -th element of a vector *x* ∈ □ ^*N*^ is denoted by *x*_*j*_ or (*x*)_*j*_. The support set or the set of indices corresponding to the nonzero elements of a vector 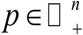 is denoted by supp(*p*).

#### Mixing matrix design

##### Parity check matrix and fixed dilution

In this section, we introduce how the participation matrix *P* and the allocation matrix *W* are designed for MHV-1 with small population size *N*, i.e., *N*=7, 15, and 31. For a prevalence rate of 1%, there can be approximately one infected sample for *N*=7, 15, and 31. From information theory, we know that the parity check matrices for Hamming codes can guarantee the correction of one error in codewords or the identification of the parity check matrix column which corresponds to the error in the codewords (23). In the context of virus testing, such parity check matrices can guarantee the identification of one positive from all the tested samples. This exactly fits our need for a small population number with 1% prevalence, and we can use such parity checking matrices as the participation matrices.

The construction of such parity check matrices can be described as follows. Suppose *P* ∈{0,1}^*n*×*N*^, then we let *N* = 2^*n*^ −1, and the columns of *P* are simply all the nonzero binary sequence of length *n*. As we consider *N*=7, 15, and 31, the corresponding participation matrices are shown in Figure 1 A-C.

**Figure 1.**
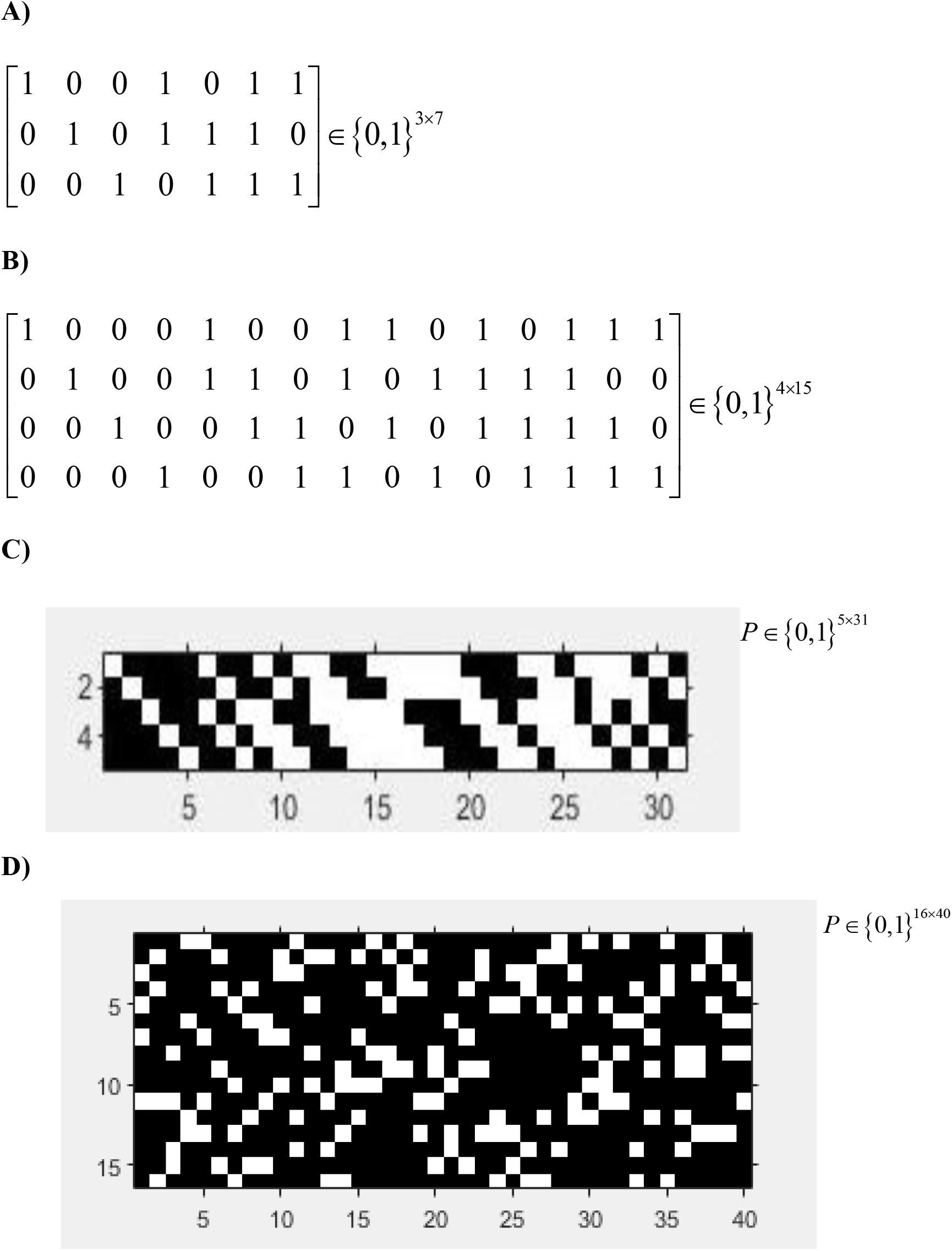
Optimized group testing mixing matrix design. **(A-C)** Hamming code parity check pooling matrix design for *N*=7, 15, and 31. **(A)** *N*=7 numerical matrix with 3 pools (3×7). **(B)** *N*=15 numerical matrix with 4 pools (4×15). **(C)** *N*=31 pixel matrix with 5 pools (5×31). **(D)** Bipartite pooling matrix design optimized for high *N* and prevalence rates. *N*=40 pixel matrix with 16 pools (16×40). **(A**,**B)** 1 indicates patient is included in the pool. 0 indicates the patient is not included in the pool. **(C**,**D)** White pixel indicates patient included in pool. Black pixel indicates patient not included in pool.

The allocation matrix should be designed in correspondence with the practical clinic procedures for mixing the samples. As for the allocation matrix *W* for MHV-1 in our laboratory experiments, since we take 5 μL from each individual sample to form the sample pool which is then concentrated to a volume of 20 μL, this implies that the virus load for an individual sample in the mixing is ¼ of its original virus load. Thus, we can design the allocation matrix as follows:

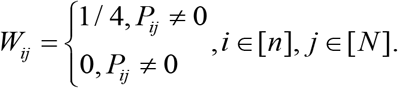

##### Bipartite graph matrix and equal partition

Though the parity check matrices of Hamming codes can be easily used as the participation matrix, it cannot scale up for high *N* or prevalence rates. This is because such parity check matrices can only guarantee the identification of one positive sample, while high *N* or prevalence rates can result in more than one positive sample in the population. Another consequence of a high *N* is the large number of nonzero elements in the participation matrix, which means high complexity during laboratory experiments. This motivates us to design participation matrices which can not only succeed in scenarios where more than 1 positive samples are present, but also have low complexity as indicated by the number of nonzero elements in the participation matrix. We propose to use the binary matrices constructed using a bipartite graph as the participation matrices (24, 25). For the COVID-19 experiments, we will use a well-designed binary matrix *P* ∈ {0,1}^16×40^ with each column having only 4 nonzero elements as shown in Figure 2.

**Figure 2.**
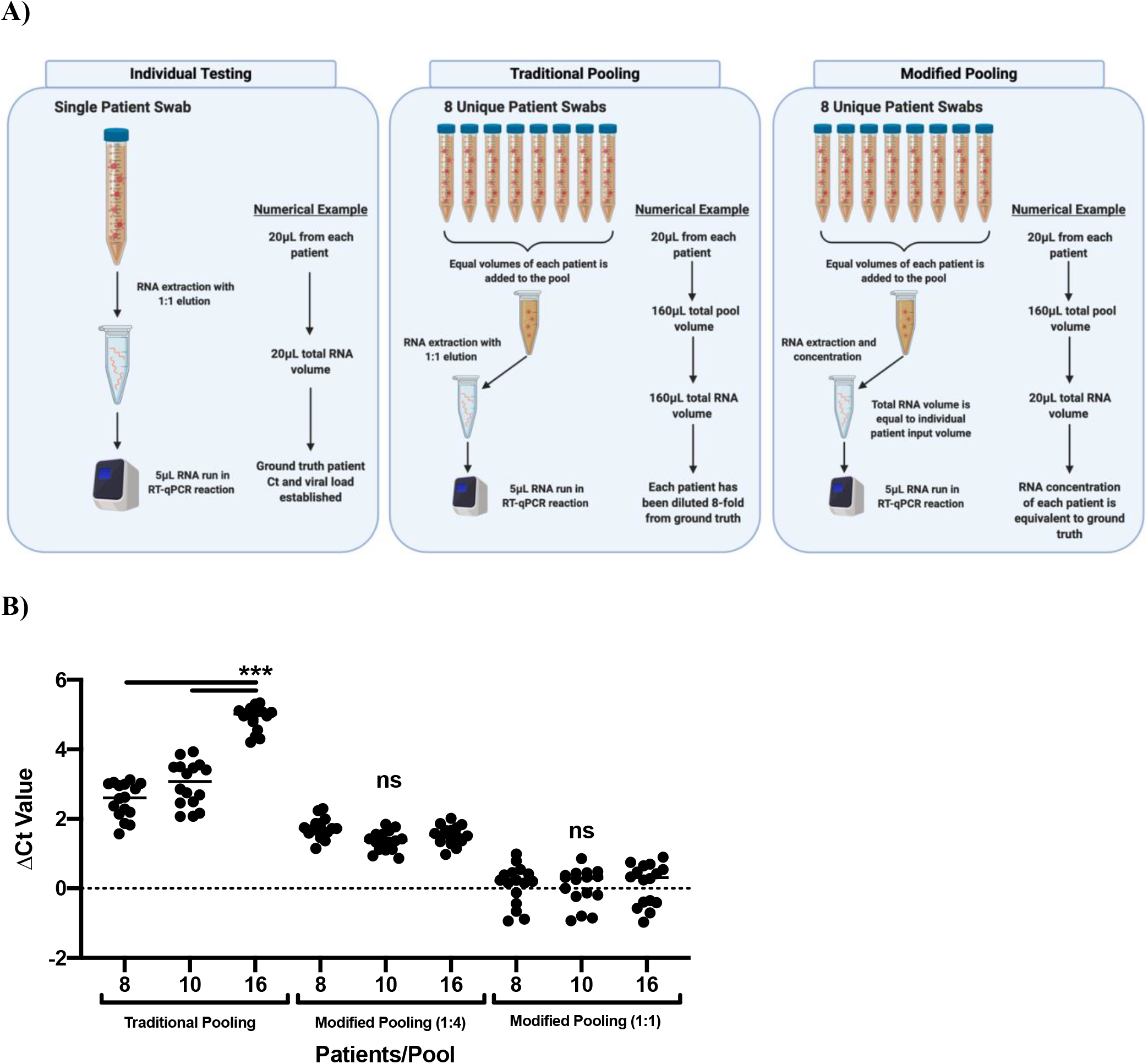
Modified pooling protocol eliminates dilution effect of group testing. (A) RNA extraction and qRT-PCR workflow in individual testing, traditional pooling (group testing), and the modified pooling protocol. Numerical examples are theoretical to display dilution effect and can be scaled to individual diagnostic testing facility protocols. (B) MHV-1 was used to generate individual samples of various viral loads (1×10^9^-1×10^2^ copy number/qRT-PCR reaction). qRT-PCR was performed on each samples to develop ground truth Ct values. Samples were then used in various pool sizes in traditional pooling and in the modified pooling protocol. Increases in sample Ct values from the ground truth values were calculated and plotted as ΔCt Value.

For SARS-CoV-2 virus testing in our laboratory experiments, since equal volumes of samples participating in a particular pool are mixed together, and we did not perform sample concentration, the virus load for each individual sample in the mixture is actually scaled down by the number of participants. Thus, the allocation matrix can be designed as:

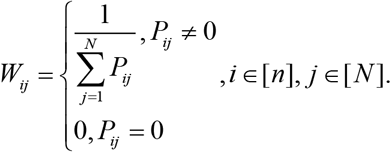

Since there is a one-to-one correspondence between the participation matrix *P* and the mixing matrix *A*, we will refer them alternatively in the subsequent sections without confusion.

##### Mixing matrix and dilution upon adaptive requests

Apart from the above pooling results with a prefixed mixing matrix, we can make requests for extra pooling results adaptively according to the decoding results at each stage. The mixing matrices used in the adaptive requests will depend on the specific decoding results, e.g., the determination of 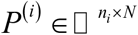 cannot be determined in advance. However, the corresponding allocation matrix will be designed according to the parity check matrix for MHV-1 and the bipartite graph matrix for SARS-CoV-2 in our laboratory experiments.

#### Sample pooling

In many group testing processes, patient samples are pooled after RNA extraction or the total pool volume dictates the RNA elution volume. In both cases, this means the fold dilution of each patient is dependent on the total number of patients within a pool. Thus, as the number of patients pooled increases, the sample become more dilute significantly increasing the probability of a false negative test result. This phenomenon has required pools to remain small, usually under 5 patients per pool (14, 15, 26). To reduce the dilution effect of pooling, a modified RNA isolation protocol was developed using TRIzol phenol/chloroform that can be more broadly applied to RNA extraction kits and automated systems such as the KingFisher (27). With this method, patient samples are pooled prior to RNA extraction. After the isopropanol precipitation and ethanol step, the pelleted RNA can be significantly concentrated by reducing the final volume of water used to solubilize the RNA thus minimizing the potential impact of sample dilution **(Fig. 2A)**.

To test the dilution effect of traditional pooling on qRT-PCR Ct results as compared to our modified RNA extraction protocol, we utilized the widely used murine coronavirus MHV-1 as a model system (28-33). Using a MATLAB-derived computational script, we pseudo-randomly generated simulated patients based on a Ct value range of 12-34 cycles. These experimental parameters were chosen from current CDC testing guidelines and growing evidence that individuals with viral loads corresponding to a Ct value of 34 and above are likely non-infectious and/or not reliable to diagnose positive patients (34-37). Additionally, in our hands, Ct values greater than 36 are generated from MHV-1 and SARS-CoV-2 qRT-PCR reactions containing ≈1-10 copies of the target gene and enter a realm where non-specific amplification and false positive rates increase.

Simulated patient samples were evaluated in qRT-PCR reactions as individuals to establish their ground truth Ct values. The samples were subsequently individually mixed with viral transport media (VTM) to generate dilutions of 8, 10, and 16-fold. The dilution was performed to simulate a situation where a single patient within a pool is positive, and consequently, the addition of other negative patient samples contributes solely to the dilution of the positive patient sample. RNA was extracted from each pool using TRIzol by either the modified RNA extraction protocol or traditional group testing. An elution volume of 20 μL was chosen to allow a 5 μL qRT-PCR test to be run in duplicate with 10 μL remaining for a retest. (**Fig. 2A)**.

As expected, samples pooled by traditional group testing exhibited a significant impact on the Ct value resulting in signal dilution (**Fig. 2B)**. However, the dilution effect was minimized or eliminated in the modified RNA extraction protocol. (**Fig. 2B)**. Importantly, the ΔCt was consistent among all pools regardless of the number of patients indicating the pool size could be significantly increased without causing further sample dilution. One issue with increasing the number of patients within a pool is the corresponding increase in the total volume of the pool. To reduce to total pool volume, we created pools by adding 5 μL of sample from each patient to the pool and eluting with 20 μL resulting in a 1:4 dilution. This approach resulted in a significantly smaller total pool volume with an average increase in Ct of 1.5 cycles with no correlation to the number of patients within the pool (**Fig. 2B)**.

These results suggest that the dilution caveat of traditional group testing can be minimized by implementing our modified extraction protocol. Patient RNA samples can also be concurrently extracted individually and banked if repeat testing is required. This approach provides a standard dilution effect that is consistent regardless of either the pool size or the volume which significantly simplifies downstream computation and decoding while reducing the chance of a false negative result.

#### Virus load decoding with success certificate

In this section, we describe a decoding algorithm which decodes each sample’s viral load from testing results of pooled samples. A unique feature of our decoding algorithm is the decoding success certificate it provides: assuming that the testing results are accurate, we are guaranteed that the decoding results are the only set of positive samples that fit the testing results.

We consider the problem of recovering a ground truth signal 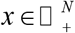 from its under-sampled measurements. Given a mixing matrix *A*∈□^*n*×*N*^ with *n* < *N*, suppose we have qualitative measurements *p* ∈{0,1}^*n*^ and qualitative measurements 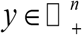 for the *n* pools which are complicated functions of 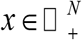, our goal is to recover 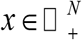 from *p* ∈{0,1}^*n*^ and 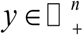. More specifically, *p* = *h*(*f* (*Ax*))where 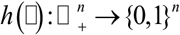, and *y* = *f* (*Ax*) where 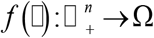 and Ω is a set of valid Ct values.

In the qRT-PCR amplification and quantification process (2) for a mixture of multiple patient samples, the quantitative relation between *b* := *Ax* and Ct value *y* can be obtained via interpolation (38). This means the function *f* (□)is the composite of the qRT-PCR amplification process and the interpolation operation. The Ct value will be compared with a threshold value *τ* preset by the authority to determine the final status of the mixture, and it varies under different scenarios. For the sake of reducing the false negative at the cost of more later tests, the technician can be conservative enough to mark positive results for mixtures although they have moderately large Ct values for which negative results can be assigned when the criterion is relaxed.

Our goal is to decode the status of *x*, i.e., positive (meaning that a sample is infected by virus) or negative (meaning that a sample is not infected by virus) status for each sample, and the amount of virus in each sample. We want to emphasize that in the virus testing practice, we will only have the Ct value data *y*, and the qualitative data *p* which is obtained from the Ct value. The *A*_*ij*_ implies whether the sample *j* participates in the *i*-th pooling test with *i* = 1, 2,…, *n* and *j* = 1, 2,…, *N*. Thus, if there is no error, a pool has positive results, i.e., *p*_*i*_ = 1 if and only if there is at least one positive element of *x* participating in the *i*-th pooling test. To achieve the above goals, we apply techniques from compressed sensing to solve it, and we end up with solving under-determined systems for *x, i*.*e*., *f* ^−1^ (*y*) = *Ax* where *f* ^−1^ is the inverse function of *f*. The problem is usually solved by min_*x*_ ║*x*║_1_, such that *f* ^−1^ (*y*) = *Ax* under the assumption that *x* is sparse (20, 22). In virus testing, the Ct value is first obtained from the qRT-PCR, and then used for interpolating the virus load *f* ^−1^ (*y*). This means the *f* ^−1^ can be treated as the interpolation procedure. We also consider min_*x*_ ║*x*║_1_, such that *f* ^−1^ (*y* + Δ*y*) = *Ax* where Δ*y* ∈ □^*n*^ characterizes the noise occurring in the measurement of Ct values.

One difference between solving under-determined systems in compressed sensing and those in the virus testing is that the values of *N* and *n* are small in the later, and large in the former. This subtle difference is critical for successful recovery, and the commonly used *L*_1_ minimization in compressed sensing may not be able to recover *x* when *N* is small. Though the accuracy outcomes are favorable when *N* is large, this is not optimal for reliability and keeping the complexity of mixing process low in clinical virus testing. **(Supplemental Fig. 1)**. Thus, in this paper we will focus on the case where *N* is small.

#### Compressed sensing decoding

In this section, we present a novel algorithm for virus decoding **(Supplemental Fig. 2A)**. Our proposed algorithm consists of three components, i.e., a support set estimation component for qualitative decoding, a quantitative decoding component which makes use of the results from the support set estimation component, and an adaptive data requesting component which asks for more testing results for improve decoding performance according to the qualitative and quantitative results.

In the support set estimation component, the goal is to give an initial estimate of the index sets of positive samples, negative samples, and samples whose status cannot be determined, respectively. We propose to solve a sequence of minimization and maximization pair for estimating an upper and a lower bound for each element of 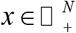, i.e., for *i* = 1, 2,…, *N*, we solve

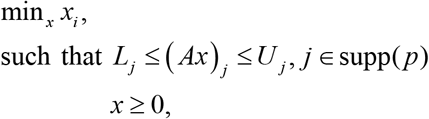

and

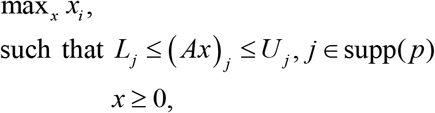

where 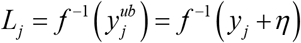 and 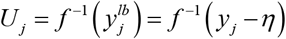 with *η* > 0 is a parameter characterizing the noise in Ct value readings. We want to emphasize that in virus testing using qRT-PCR, a larger Ct value corresponds to a smaller virus load (2). After we get the lower (upper) bound estimates 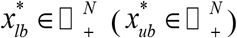, we compare each of its element with a upper bound virus load threshold parameter *ε*_νlub_ ∈ □ _+_ (*ε*_νl*lb*_ ∈ □ _+_). If 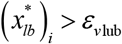 or 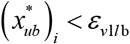, we claim the *i* -th sample of *x* must be positive or negative. By repeating the comparison for each *i* ∈[*N*], we can obtain index sets *Pos* and *Neg* which are the index sets of samples which must be positive and negative, respectively. Finally, the index set of samples whose status cannot be determined can be obtained as *U* := [*N*]\ (*Pos* ∪ *Neg*). The above algorithm is presented in Algorithm 2 **(Supplemental Fig. 2B)**.

The set estimates *Pos, Neg,U* are then exploited in the quantitative decoding component whose core is an exhaustive search algorithm. For the exhaustive search component, we solve a weighted least square for each possible cardinality *k* ∈{1, 2,…, |*U* |} and for each possible support set *K* ⊆ *U* with cardinality |*K*| = *k*, i.e.,

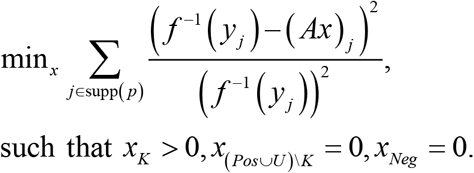

The main idea is to estimate a sample virus load 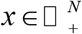 such that the deviation between the estimated pool virus load (*Ax*) _*j*_ and the corresponding interpolated pool virus load *f* ^−1^ (*y*_*j*_) is minimized. Due to the wide range that the sample virus load can reside, i.e., from 10^−6^ to 10^6^, we normalize the deviation via a scaling factor 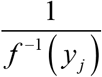 The algorithm is presented in supplemental files. Usually in practice, the combinatorial characteristics of exhaustive search can bring high computational complexity and high accuracy. In our virus testing problem, due to the small size of the problem, the exhaustive search can be a good option. Besides, the support set estimation component can be used to further reduce the size of the combinatorial problem. Another trick we use to reduce the computational complexity is that we try to find the sparsest solution. This is achieved by finding the solution with the smallest support set such that the misfit between the estimated Ct value and the measured Ct value is smaller than a given tolerance for all the observed positive pools.

In the data adaptive requesting component, based on the results from the support set estimation and the quantitative decoding components, we design new pooling strategies for pooling samples. The extra pooled testing results are obtained using individual samples whose status and virus load cannot be determined by previous pooled testing results. The mixing matrices for pooling the undetermined individual samples can be case-specific in practice. The algorithm is presented in Algorithm 3 **(Supplemental Fig. 2C)**. Usually in practice, the combinatorial characteristics of exhaustive search can bring high computational complexity though it can achieve high accuracy for estimating *x*. In our virus testing problem, due to the small size of the problem, the exhaustive search can be a good option. Besides, the Algorithm 2 can be used to further reduce the size of the combinatorial problem. Another trick we use to reduce the computational complexity is that we try to find the sparsest solution. This is achieved by finding the solution with the smallest support set such that the misfit between the estimated Ct value and the measured Ct value is smaller than a given tolerance Δ*y* for all the observed positive pools.

## Results

To demonstrate proof of concept, we began our initial experiments with the model coronavirus MHV-1 testing a range of experimental parameters (28-33). As in the pooling dilution effect experiments, a MATLAB-based script was used to generate pseudorandom experimental parameters based on *N* total samples with a prevalence rate of 1-10%. Samples were mixed together to form *n* different pools according to the participation matrix in Figure 1. Total RNA was extracted from the generated pools utilizing our 1:4 modified pooling technique **(Fig. 2A)**. Total RNA isolated from sample pools was then amplified via qRT-PCR to generate a numerical readout of cycle threshold values. To avoid accidental errors, for every group of *N* samples and a given mixing matrix *A* ∈ □ ^*n×N*^ (here *n* is just *n*_1_ in Algorithm 1) experiments were duplicated.

In one of our experiments, pools 1, 2, 4, and 5 returned Ct values within the bounds to be considered positive **(Table 1)**. With this information alone, Algorithm 2 can decode the samples with *Neg* ={3,6,8,9,12,13,14,15,16, 20, 21, 23, 24, 25, 27, 29} as negative, and the rest of the samples are undetermined. This means *U* = {1, 2, 4,5, 7,10,11,17,18,19, 22, 26, 28,30,31}, and *Pos* = Ø **(Supplementary Table 1)**. These sets are consistent with the virus load decoded by exhaustive search in which the samples decoded by Algorithm 2 as negative indeed have almost zero virus load, while those which are decoded as undetermined have virus loads which are neither too big nor too small to be considered negative. However, from the decoding results from Algorithm 3, we can see that apart from giving zero estimate for the virus load of samples specified by *Neg*, it also estimates all samples from *U*, except sample 17, to have zero virus load. This can be validated with request for one extra pooling test involving all the samples in *U* except 17.

**Table 1.** *N*=31 MHV-1 pooled testing qRT-PCR results.

| **Table 1. *N*=31 MHV-1 pooled testing qRT-PCR results** |  |  |  |
| --- | --- | --- | --- |
| **Pool #** | **Status** | **Ct Duplicate 1** | **Ct Duplicate 2** |
| Pool 1 | Pos | 35.068 | 34.234 |
| Pool 2 | Pos | 35.107 | 34.526 |
| Pool 3 | Neg | NA | NA |
| Pool 4 | Pos | 35.021 | 34.697 |
| Pool 5 | Pos | 34.031 | 34.123 |

After initial pooling and decoding, further pooling for confirmation testing may be required. We will refer to the matrix in Figure 1C as *P*^(1)^. From our decoding result, we request an additional pooling test (*P*^(2)^ ) since not all sample infection statuses can be determined with 100% certainty. Thus, we designed the mixing matrix which pools all the samples that are highly likely false positive **(Supplementary Fig. 3)**. Viral loads which are very small in magnitude can be due to numerical error, and we can simply treat it as 0.

Overall, the infection status of 1325 unique experimentally generated samples were determined with individual experimental prevalence rates ranging from 0-14.3% **(Table 2)**. After a single round of testing, the infection status of 97.4% of all samples was established with 100% certainty. One subsequent round of verification testing identified the infection status of 98.9% samples with full certainty and 15 remaining samples which required further testing to determine infection status with full certainty.

**Table 2.** Human COVID-19 sample pooled testing qRT-PCR results.

| **Pool #** | **Status** | **Ct Duplicate 1** | **Ct Duplicate 2** |
| --- | --- | --- | --- |
| Pool 1 | Pos | 33.92 | 32.961 |
| Pool 2 | Pos | 20.68 | 20.909 |
| Pool 3 | Pos | 34.065 | 36.231 |
| Pool 4 | Pos | 26.562 | 27.051 |
| Pool 5 | Neg | NA | NA |
| Pool 6 | Pos | 26.719 | 26.864 |
| Pool 7 | Pos | 19.977 | 20.386 |
| Pool 8 | Pos | 27.063 | 27.756 |
| Pool 9 | Neg | NA | NA |
| Pool 10 | Pos | 20.636 | 20.945 |
| Pool 11 | Pos | 27.574 | 27.266 |
| Pool 12 | Pos | 20.196 | 20.925 |
| Pool 13 | Neg | NA | NA |
| Pool 14 | Pos | 32.336 | 32.185 |
| Pool 15 | Neg | NA | NA |
| Pool 16 | Pos | 33.133 | 32.97 |

In total, 322 tests were required to identify all positive samples within the population of 1325 total samples. This resulted in a 75.6% reduction in the total number of tests required as compared to individualized testing. These experiments were repeated with similar parameters and results bringing the total number of experimentally generated samples tested to 2650.

To validate our pooling and detection system, we obtained human patient RNA samples from the University of Iowa diagnostic testing laboratory. Samples were provided as extracted RNA, thus our modified RNA extraction protocol was not utilized and samples were mixed using traditional pooling **(Fig. 2A)**. An optimized participation matrix was generated to reflect the expected dilution effect **(Fig 1D)**. Experimental parameters were pseudo-randomly generated as previously described with a total *N* of 40 patients and a set prevalence rate of 10%. The pooling results for one of two independent experiments is presented in table 3. For both of the two runs, we requested extra pooling results for decoding, and thus required the generation of an additional mixing matrix **(Supplementary Fig. 4)**. Additional pooling results and individual patient viral loads is shown in supplementary tables 2 and 3.

**Table 3.**
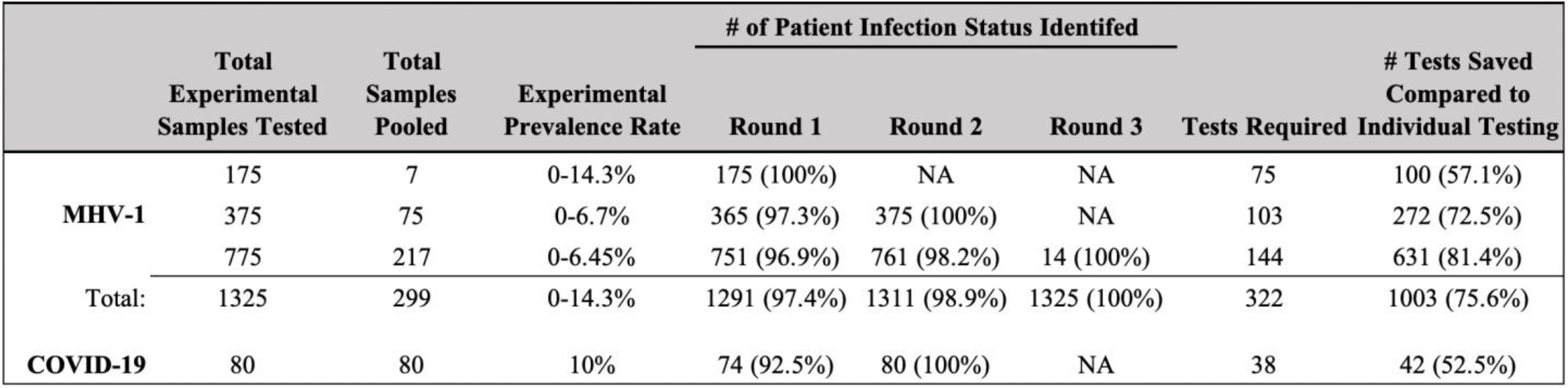
Compressed sensing decoded pooled testing significantly decreases the number of tests required to identify infected patients.

After one round of testing and compressed sensing decoding, 2 patients were identified and confirmed as positive and 72 were confirmed as negative leaving 6 patients as likely positive. Two subsequent pools and four individual confirmation tests provided adequate data points to determine the infection status of all patients with 100% certainty. 32 tests were required to determine the infection status of 92.5% of all patients. Additional confirmatory testing brought the total tests performed to screen 80 patients to 38. This is a 52.5% reduction in the number of tests needed as compared to current individual testing **(Table 2)**.

## Discussion

Together, our experimental data provides proof of concept and validates our compressed sensing pooling system as an effective and reproducible method to greatly increase COVID-19 testing capacity while simultaneously providing more diagnostic information by determining patient viral load. Using our novel testing approach, we were able to identify positive samples with extreme accuracy at prevalence rates at 10% or higher in both an MHV-1 coronavirus model system and human COVID-19 patient samples. This required approximately one third as many tests as would be needed with current individual testing procedures.

Pooled testing is an effective approach to increase testing capacity and allow widespread screening to occur and has been implemented with limited success during the COVID-19 pandemic (9-14, 26, 39-44). However, current pooled testing efforts lose efficacy and precision at real world prevalence rates and ultimately require substantial additional confirmation testing. In 2020 for the first time in the field, we proposed to use compressed sensing techniques for quantitative virus testing with high prevalence, and computational experiments validated the effectiveness of our method (42). Others such as Ghosh et al. and Shental et al., showed the superiority of compressed sensing virus testing technology using a non-adaptive approach though their method could only succeed at low prevalence, e.g., less than 10% (43, 44). In contrast, our current work uses an adaptive approach and can succeed at prevalence rates greater than 10% and utilizes a success certificate to ensure results are accurate (**Section S1.3**). Additionally, one major caveat of pooled testing is sample dilution and the increase of false negatives. To eliminate the pooling dilution effect, we utilized a modified RNA extraction protocol which differs from current clinical diagnostic lab procedures by simply concentrating the RNA to a set volume regardless of the patient input number **(Fig. 2A)**. This standardizes the dilution to an expected and reproducible ΔCt from the ground truth value that does not change if the number of patients within a pool increases **(Fig. 2B)**. This protocol alone removes the risk of samples with low levels of virus being diluted in a pool and being read as a false negative.

Our approach demonstrates an effective process to combat testing bottlenecks for future pandemics. Many clinical testing labs currently utilize automated RNA extraction systems in which parameters can be changed to fit our new protocols. Additionally, we have created a beta decoding software in which qPCR data can be entered and the program will decode the data, identify positive patients, and generate additional pools for further testing, if needed, all automatically (software code available upon request). Most importantly, the application of our testing method is broad and can be applied to many testing applications within medicine and beyond such as serum antibody testing, drug screening, avian influenza surveillance, water contamination testing, etc. Our application of compressed sensing is perfectly positioned for testing applications such as these as they are sparse by nature and require accurate results from many data points.

The emergence of new pathogens and deadly variants is ongoing and will continue to be a significant threat to public health and humanity as a whole (4-8). Implementing a highly accurate pooled testing procedure is absolutely critical to mitigating the spread of deadly viral pandemics such as COVID-19, thus saving lives and decreasing the economic destruction from high mortality rates and widespread quarantines. Our use of compressed sensing in pooled COVID-19 testing demonstrated high sensitivity in experimental infection models with the model coronavirus MHV-1, as well as with primary human COVID-19 samples. The utilization of compressed sensing theory in signal analysis is well established, but its use in the testing of physical specimens has the potential to revolutionize how we provide accurate results when testing extremely large numbers of samples. This will position healthcare professionals to rapidly respond to future pandemics by identifying infected individuals early, minimizing spread, and thus saving lives.

## Materials and Methods

### Generation of experimental parameters and positive MHV-1 samples

We used a computer script to generate pseudorandom viral loads for each of *N* individual samples based on an average prevalence rate of 5%, and positive patient Ct values in the range 12-34. The MHV-1 standard curve was used to plot the generated sample Ct value (X) and interpolate the dilution of MHV-1 virus stock (Y) required. According to these estimates *Y*, MHV-1 was diluted in viral transport media as in the CDC-approved nasopharyngeal swab collection protocol. (34, 36).

### MHV-1 sample pooling

5-20 μL of generated MHV-1 samples were pooled together in equal volumes on ice as designated by the appropriate mixing matrix. Negative samples were added as sterile viral transport media.

### Human patient sample pooling

Human samples that were to be discarded were supplied as extracted RNA in 96-well plates from the University of Iowa Diagnostic Testing Lab. Patients were identified as positive or negative with no information on Ct number, viral load, or any patient identifiable information. 5 μL of patient samples were pooled together in equal volumes on ice as designated by the appropriate mixing matrix. The University of Iowa determined that this project did not meet the regulatory definition of human subjects research and therefore IRB approval was not required.

### Isolation of viral RNA

Viral RNA was extracted via a modified TRIzol phenol/chloroform extraction protocol and can be scaled as needed **(Fig 2)**. A patient pool of 20 μL total volume was mixed with 200 μL TRIzol. The sample was vortexed for 10 sec and incubated for 5 min at room temperature (RT). 40 μL of chloroform was added, vortexed for 10 sec, and incubated for 5 min at RT. The mixture was centrifuged at 12,000 x g for 10 min at 4°C. 100 μL of the upper aqueous layer was transferred to a sterile 1.5 mL tube. 100 μL of isopropanol supplemented with 2 μg glycogen was added, vortexed for 10 sec, and incubated for 5 min at RT. The pellet was mixed with 180 μL of 75% ethanol and resuspended by gentle inversion and centrifuged at 14,000 x g for 10 min at RT. The supernatant was aspirated and the pellet was air dried for 10 min in a sterile laminar flow hood. The RNA pellet was resuspended in 20 μL of RNAse-free diethyl pyrocarbonate-treated H_2_O and incubated at 55°C for 5 min.

### qRT-PCR

5μL of patient pools and samples were mixed with the GoTaq qRT-PCR master mix (Promega) and ran in duplicate on a QuanStudio 3 thermocycler via the FAST qRT-PCR protocol as recommended by the CDC (36). An MHV-1 virus stock or SARS-Cov-2 S protein containing plasmid of known concentrations were used to generate a standard curve consisting of seven to ten 10-fold serial dilutions. The resulting amplification curves were analyzed with AppliedBiosystems Design and Analysis 2.4.

### Compressed Sensing Decoding

An optimization algorithm leveraging the non-negativity of viral loads was used to give an upper and lower bound on the viral load for each sample. If the lower bound for a sample’s viral load is not zero, we are sure that that sample is positive; if the upper bound for a sample’s viral load is equal to 0, we are sure that that sample is negative. This identifies samples which are either definitely positive or definitely negative. For the samples with ambiguous infection statuses, we perform exhaustive search for the smallest set of positive samples (namely sparsest solution, having the smallest number of positive samples) fitting the observed viral loads of these pools. The remaining samples were mixed together into a pooled sample to confirm that they are indeed negative: if this pooled sample comes back positive, further testing will be necessary, but this is statistically unlikely.

## Supporting information

Ethical approval Letter

## Data Availability

The implementation of our system and the data used for conducting the experiments can be provided once requested.

## Author contributions

Conceptualization: RM, XW, SMV, WX

Methodology: KAW, JY, WX, MC

Investigation: KAW, JY

Funding acquisition: SMV, WX

Supervision: SMV, WX,

Writing – original draft: KAW

Writing – review & editing: KAW, JY, WX, SMV

## Data and materials availability

All data, code, and materials used in this study are available upon request.

