## Supplemental data for "Use of compressed sensing to expedite high-throughput diagnostic testing for COVID-19 and beyond"

**Fig. S1. Compressed sensing accuracy increases with *N*.**

Random test simulation to assess the performance of compressed sensing at low and high *N*. A Bernoulli random matrix $A \in \{0,1\}^{n \times N}$ with $\Pr(A_{ij}=0)=\Pr(A_{ij}=1)=0.5$ is used for both cases. We take $n = \mathrm{round}(0.3 * N)$, and the $x$ is generated uniformly from $[0,100]^N$ with sparsity $\mathrm{round}(0.05 * N)$. The horizontal axis is the index element of x. The vertical axis is the value of the element. **(A)** *N*=10. **(B)** *N*=100

Field Code Changed
Field Code Changed
Field Code Changed
Field Code Changed
Field Code Changed

**A)**

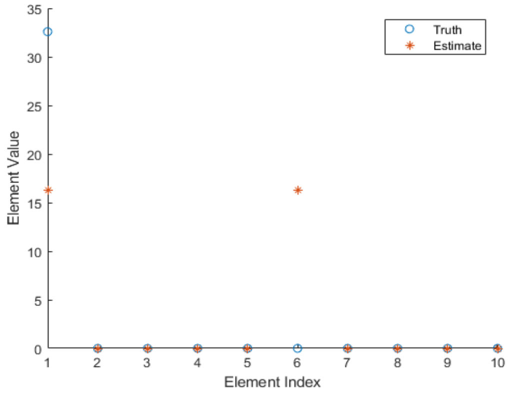


**B)**

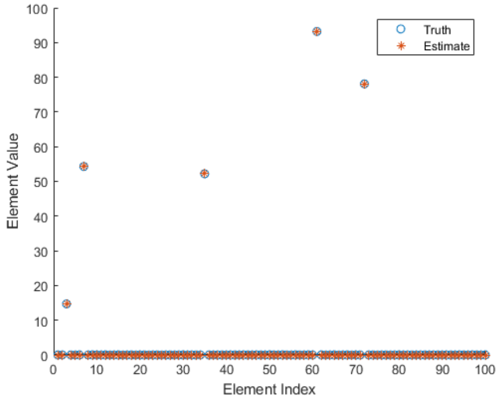

**Fig. S2. Compressed sensing decoding algorithms.**

**(A)** Algorithm 1 virus decoding. **(B)** Algorithm 2 support estimation. **(C)** Algorithm 3 exhaustive search

**(A)**

---

**Algorithm 1** Virus decoding algorithm

---

**Require:** Virus load lower bound $\epsilon_{vllb} \in \mathbb{R}_+$, virus load upper bound $\epsilon_{vlub} \in \mathbb{R}_+$, $c_t$ value tolerance $\Delta y$, $c_t$ value noise level $\eta \in \mathbb{R}_+$, parameter for avoiding numeric error $\epsilon_{num} \in \mathbb{R}_+$, pooling results $p^{(1)} \in \{0,1\}^{n_1}$, $c_t$ value reading $y^{(1)} \in \mathbb{R}_+^{n_1}$, mixing matrix $A^{(1)} \in \mathbb{R}_+^{n_1 \times N}$

$i = 1$
Define $A^* = A^{(1)} \in \mathbb{R}_+^{n_1 \times N}$
Define $y^* = y^{(1)} \in \mathbb{R}_+^{n_1}$
Define $p^* = p^{(1)} \in \{0,1\}_+^{n_1}$
Call Algorithm 2 with $A^*, p^*, y^*$ to get estimated index sets $\mathcal{P}, \mathcal{N}, \mathcal{U}$ of positive samples, negative samples, and samples whose status cannot be determined respectively
Define $\mathcal{U}^* = \mathcal{U}$
**while** $\mathcal{U}^* \neq \emptyset$ **do**
  $i \leftarrow i + 1$
  Request pooling results $p^{(i)} \in \{0,1\}^{n_i}$, $c_t$ value reading $y^{(i)} \in \mathbb{R}_+^{n_i}$, mixing matrix $A^{(i)} \in \mathbb{R}_+^{n_i \times N}$ according to $x^*$
  $A^* \leftarrow \begin{bmatrix} A^* \\ A^{(i)} \end{bmatrix} \in \mathbb{R}_+^{\sum_{j=1}^{i} n_i \times N}$
  $y^* \leftarrow \begin{bmatrix} y^* \\ y^{(i)} \end{bmatrix} \in \mathbb{R}^{\sum_{j=1}^{i} n_i}$
  $p^* \leftarrow \begin{bmatrix} p^* \\ p^{(i)} \end{bmatrix} \in \{0,1\}^{\sum_{j=1}^{i} n_i}$
  Call Algorithm 2 with $A^*, p^*, y^*$ to get estimated index sets $\mathcal{P}, \mathcal{N}, \mathcal{U}$ of positive samples, negative samples, and samples whose status cannot be determined respectively
  Define $\mathcal{U}^* = \mathcal{U}$
**end while**
Define $\mathcal{S} = \mathrm{supp}(p)$
Call Algorithm 3 with $A^*, p^*, y^*$ to get $x^* \in \mathbb{R}_+^N$
Return $x^* \in \mathbb{R}_+^N, \mathcal{P}, \mathcal{U}, \mathcal{N}$

---

**B)**

**Algorithm 2** Support estimation

**Require:** Mixing matrix $A \in \mathbb{R}_+^{n \times N}$, $c_t$ value $c_t \in \mathbb{R}_+^n$, pooling results $p \in 0, 1^n$, noise level $\eta \in \mathbb{R}_+$, virus load lower bound $\epsilon_{vllb}$, and virus load upper bound $\epsilon_{vlub}$

**for** $i \in [N]$ **do**

  Solve

$$x_{lb} = \arg\min_{x \in C} x_i$$

  and

$$x_{ub} = \arg\max_{x \in C} x_i$$

  where $C := \{x \in \mathbb{R}_+^n : L_j \le Ax \le U_j, \forall j \in \text{supp}(p)\}$, $L_j = f^{-1}(y_j^{ub}) = f^{-1}(y_j + \eta)$, and $U_j = f^{-1}(y_j^{lb}) = f^{-1}(y_j - \eta)$ with constant parameter $\eta > 0$

**end for**

Define estimated index set of negative samples $\mathcal{N} = \emptyset$

Define estimated index set of positive samples $\mathcal{P} = \emptyset$

Define estimated index set of samples whose status cannot be determined $\mathcal{U} = \emptyset$

**for** $i \in [N]$ **do**

  **if** $(x_{ub})_i < \epsilon_{vllb}$ **then**

    Add $i$ to $\mathcal{N}$

  **else if** $(x_{lb})_i > \epsilon_{vlub}$ **then**

    Add $i$ to $\mathcal{P}$

  **end if**

**end for**

$\mathcal{U} = [N] \setminus (\mathcal{P} \cup \mathcal{N})$

Return $\mathcal{P}$,$\mathcal{N}$,$\mathcal{U}$

**C)**

**Algorithm 3** Exhaustive search

**Require:** Pooling results $p \in \{0,1\}^n$, $c_t$ value $y \in \mathbb{R}_+^n$, $c_t$ value tolerance $\Delta y \in \mathbb{R}_+$, mixing matrix $A \in \mathbb{R}_+^{n \times N}$, index sets $\mathcal{P}, \mathcal{N}, \mathcal{U}$ from Algorithm 2

Define maximal sparsity $MaxSpst = |\mathcal{P} \cup \mathcal{U}|$
Define support set $\mathcal{S} = \mathrm{supp}(p)$
Define $obj^* = \infty$ and $x^* = 0 \in \mathbb{R}_+^N$
Define early stop flag $EarlyStop = 0$
**for** $k = 1, 2, \cdots, MaxSpst$ **do**
  **for** every subset $\mathcal{K} \subset (\mathcal{P} \cup \mathcal{U})$ with cardinality $k$ **do**
    Solve the (8) to get the optimal objective function value $obj$ and optimal solution $x$

$$obj = \min_x \sum_{i \in \mathcal{S}} \frac{1}{(f^{-1}(y)_i)^2} \left((Ax)_i - f^{-1}(y)_i\right)^2,$$
$$\text{s.t. } x_{\mathcal{K}} \geq 0, x_{[N] \setminus \mathcal{K}} = 0$$

    **if** $obj < obj^*$ **then**
      $obj^* = obj$
      $x^* = x$
      Define $\hat{y} \in \mathbb{R}_+^n$ with $\hat{y}_{\mathcal{S}} = f(A_{\mathcal{S}} x^*)$ and $\hat{y}_{[n] \setminus \mathcal{S}} = NA$
      **if** $|\hat{y}_i - y_i| < \Delta y, \forall i \in \mathcal{S}$ **then**
        $EarlyStop = 1$
        Break
      **end if**
    **end if**
  **end for**
  **if** $EarlyStop = 1$ **then**
    Break
  **end if**
**end for**
Return $x^*$

**Fig. S3. Adaptive request pooling matrix.**

Pooling matrix designed for additional testing requests. 1 indicates sample is included in the pool. 0 indicates the sample is not included in the pool.

$$P^{(2)} = \begin{bmatrix} 1 & 1 & 0 & 1 & 1 & 0 & 1 & 0 & 0 & 1 & 1 & 0 & 0 & 0 & 0 & 0 & 0 & 1 & 1 & 0 & 0 & 1 & 0 & 0 & 0 & 1 & 0 & 1 & 0 & 1 & 1 \end{bmatrix} \in \sim^{1\times 31}$$

**Fig. S4. Human COVID-19 additional testing pooling matrix.**

Pooling matrix designed for additional testing requests in human COVID-19 samples. $N$=40 (3x40). 1 indicates patient is included in the pool. 0 indicates the patient is not included in the pool.

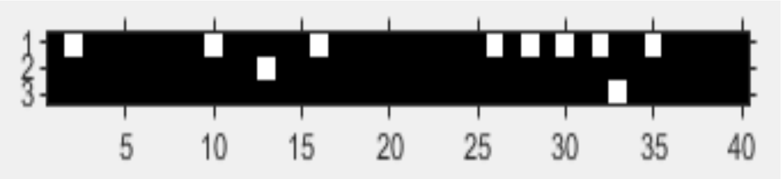

| Supplementary Table 1. MHV-1 individual sample infection status after one round of testing | | | | |
|---|---|---|---|---|
| | | Sample Viral Load (ng/mL) | | |
| **Patient** | **Status** | **Lower Bound** | **Upper Bound** | **Estimated Viral Load** |
| 1 | Undetermined | 2.01E-10 | 1.20E-03 | 0.00E+00 |
| 2 | Undetermined | 2.54E-10 | 1.17E-03 | 0.00E+00 |
| 3 | Negative | 5.68E-13 | 1.09E-12 | 0.00E+00 |
| 4 | Undetermined | 2.05E-10 | 1.23E-03 | 0.00E+00 |
| 5 | Undetermined | 1.34E-10 | 2.20E-03 | 0.00E+00 |
| 6 | Negative | 7.49E-13 | 6.88E-13 | 0.00E+00 |
| 7 | Undetermined | 1.82E-10 | 1.17E-03 | 0.00E+00 |
| 8 | Negative | 6.18E-13 | 7.53E-13 | 0.00E+00 |
| 9 | Negative | 7.55E-13 | 7.64E-13 | 0.00E+00 |
| 10 | Undetermined | 8.62E-11 | 1.17E-03 | 0.00E+00 |
| 11 | Undetermined | 2.09E-10 | 1.20E-03 | 0.00E+00 |
| 12 | Negative | 7.62E-13 | 7.64E-13 | 0.00E+00 |
| 13 | Negative | 7.60E-13 | 7.66E-13 | 0.00E+00 |
| 14 | Negative | 7.30E-13 | 7.54E-13 | 0.00E+00 |
| 15 | Negative | 7.26E-13 | 9.18E-13 | 0.00E+00 |
| 16 | Negative | 1.21E-12 | 6.33E-13 | 0.00E+00 |
| 17 | Undetermined | 9.50E-11 | 1.17E-03 | 1.24E-03 |
| 18 | Undetermined | 8.83E-11 | 1.17E-03 | 0.00E+00 |
| 19 | Undetermined | 1.95E-10 | 1.17E-03 | 0.00E+00 |
| 20 | Negative | 7.54E-13 | 6.91E-13 | 0.00E+00 |
| 21 | Negative | 7.53E-13 | 6.88E-13 | 0.00E+00 |
| 22 | Undetermined | 1.96E-10 | 1.23E-03 | 0.00E+00 |
| 23 | Negative | 7.33E-13 | 7.48E-13 | 0.00E+00 |
| 24 | Negative | 7.53E-13 | 9.58E-13 | 0.00E+00 |
| 25 | Negative | 7.28E-13 | 9.24E-13 | 0.00E+00 |
| 26 | Undetermined | 8.57E-11 | 1.20E-03 | 0.00E+00 |
| 27 | Negative | 7.29E-13 | 9.21E-13 | 0.00E+00 |
| 28 | Undetermined | 8.22E-11 | 1.17E-03 | 0.00E+00 |
| 29 | Negative | 7.38E-13 | 7.48E-13 | 0.00E+00 |
| 30 | Undetermined | 1.86E-10 | 1.20E-03 | 0.00E+00 |
| 31 | Undetermined | 2.22E-10 | 1.17E-03 | 0.00E+00 |

| **Supplementary Table 2. Human COVID-19 sample second round pooling qRT-PCR results** | | | |
|---|---|---|---|
| **Pool #** | **Status** | **Ct Duplicate 1** | **Ct Duplicate 2** |
| Pool 1 | Neg | NA | NA |
| Pool 2 | Pos | 29.592 | 30.125 |
| Pool 3 | Neg | NA | NA |

| Supplementary Table 3. Human COVID-19 individual patient infection status results | | | | |
|---|---|---|---|---|
| | | Sample Viral Load (ng/mL) | | |
| Patient | Status | Lower Bound | Upper Bound | Estimated Viral Load |
| 1 | Negative | 2.29E-12 | 1.85E-12 | 0.00E+00 |
| 2 | Negative | 1.34E-12 | 9.90E-13 | 0.00E+00 |
| 3 | Negative | 2.13E-12 | 2.46E-12 | 0.00E+00 |
| 4 | Negative | 2.18E-12 | 1.23E-13 | 0.00E+00 |
| 5 | Negative | 1.54E-12 | 2.46E-12 | 0.00E+00 |
| 6 | Negative | 1.03E-12 | 1.13E-12 | 0.00E+00 |
| 7 | Negative | 7.04E-13 | 6.42E-13 | 0.00E+00 |
| 8 | Negative | 2.15E-12 | 2.32E-12 | 0.00E+00 |
| 9 | Negative | 1.78E-12 | 2.52E-12 | 0.00E+00 |
| 10 | Negative | 9.34E-13 | 1.28E-12 | 0.00E+00 |
| 11 | Positive | 2.35E+01 | 3.95E+01 | 2.90E+02 |
| 12 | Negative | 1.50E-12 | 1.22E-13 | 0.00E+00 |
| 13 | Positive | 2.57E+02 | 3.56E+02 | 1.01E+03 |
| 14 | Negative | 9.13E-13 | 7.00E-13 | 0.00E+00 |
| 15 | Positive | 2.37E+05 | 2.04E+06 | 5.79E+06 |
| 16 | Negative | 1.13E-12 | 1.25E-12 | 0.00E+00 |
| 17 | Negative | 9.23E-13 | 1.00E-12 | 0.00E+00 |
| 18 | Negative | 3.36E-12 | 2.62E-12 | 0.00E+00 |
| 19 | Negative | 1.88E-12 | 1.58E-12 | 0.00E+00 |
| 20 | Negative | 9.38E-13 | 1.28E-12 | 0.00E+00 |
| 21 | Negative | 1.39E-12 | 2.19E-12 | 0.00E+00 |
| 22 | Negative | 1.03E-12 | 1.10E-12 | 0.00E+00 |
| 23 | Negative | 7.97E-13 | 8.96E-13 | 0.00E+00 |
| 24 | Negative | 6.23E-13 | 7.14E-13 | 0.00E+00 |
| 25 | Negative | 3.59E-13 | 2.52E-13 | 0.00E+00 |
| 26 | Negative | 8.55E-13 | 1.38E-12 | 0.00E+00 |
| 27 | Negative | 2.01E-12 | 2.14E-12 | 0.00E+00 |
| 28 | Negative | 1.21E-12 | 9.71E-14 | 0.00E+00 |
| 29 | Negative | 2.30E-12 | 1.72E-12 | 0.00E+00 |
| 30 | Negative | 1.07E-12 | 1.27E-12 | 0.00E+00 |
| 31 | Negative | 9.47E-13 | 1.03E-12 | 0.00E+00 |
| 32 | Negative | 1.20E-12 | 9.82E-13 | 0.00E+00 |
| 33 | Negative | -2.29E-16 | 1.85E-16 | 0.00E+00 |
| 34 | Negative | 9.52E-13 | 1.02E-12 | 0.00E+00 |
| 35 | Negative | 1.10E-12 | 1.25E-12 | 0.00E+00 |
| 36 | Negative | 2.83E-12 | 2.80E-12 | 0.00E+00 |
| 37 | Negative | 7.93E-13 | 9.05E-13 | 0.00E+00 |
| 38 | Negative | 5.44E-14 | 7.80E-13 | 0.00E+00 |
| 39 | Negative | 1.23E-12 | 2.00E-12 | 0.00E+00 |
| 40 | Positive | 2.21E+03 | 1.62E+04 | 6.48E+04 |
