## Supplementary material for "Use of compressed sensing to expedite high-throughput diagnostic testing for COVID-19 and beyond": Ethical approval Letter

THE UNIVERSITY OF IOWA

**Human Subjects Office/**
**Institutional Review Board (IRB)**
105 Hardin Library for the Health Sciences
600 Newton Road
Iowa City, Iowa 52242-1098

http://research.uiowa.edu/hso

July 16, 2021

TO: Steven Varga

FROM: Catherine Woodman
IRB Chair

RE: Not Human Subjects Research Determination

I have reviewed the information submitted with your project titled [IRB#202103412][Error-Correcting Quantitative Pool Testing for COVID-19].

I have determined that the project described in the application *does not* meet the regulatory definition of human subjects research and does not require review by the IRB, because the UI research team will not interact with subjects and will be provided only completely deidentified specimens for analysis.

**If in the future you undertake similar activities, I encourage you to start with submission of a Human Subjects Research Determination (HSRD) form in the HawkIRB eIRB system.**

We appreciate your care in submitting this application to the IRB for review. If the parameters outlined within this Human Subjects Research application request change, re review and/or subsequent IRB review may be required.

Please don't hesitate to contact me if you have any questions. The Human Subjects Office can be reached via phone (319)-335-6564 or.
